# Safety and effects of acetylated and butyrylated high amylose maize starch in recently diagnosed youths with type 1 diabetes; a Pilot Study

**DOI:** 10.1101/2024.05.17.24307489

**Authors:** Heba M Ismail, Jianyun Liu, Michael Netherland, Carmella Evans-Molina, Linda A DiMeglio

**Affiliations:** Department of Pediatrics, Indiana University School of Medicine, Indianapolis, IN, USA; Department of Surgery, Indiana University School of Medicine, Indianapolis, IN, USA; EzBiome, Inc., Gaithersburg, MD, USA

**Keywords:** HAMS-AB, dietary intervention, type 1 diabetes (T1D), Beta cell function, mixed meal tolerance test (MMTT), glucose regulation, C-peptide levels, glycemic control, metabolomics, SCFAs

## Abstract

Acetylated and butyrylated high amylose starch (HAMS-AB) is a prebiotic shown to be effective in type 1 diabetes (T1D) prevention in mouse models and is safe in adults with established T1D. HAMS-AB alters the gut microbiome profile with increased bacterial fermenters that produce short chain fatty acids (SCFAs) with anti-inflammatory and immune-modulatory effects. We performed a pilot study using a cross-over design to assess the safety and efficacy of 4 weeks of oral HAMS-AB consumption by recently diagnosed (< 2 years of diagnosis) youths with T1D. Seven individuals completed the study. The mean±SD age was 15.0±1.2 years, diabetes duration 19.5±6.3 months, 5/7 were female and 4/7 were White, all with a BMI of < 85^th^%. The prebiotic was safe. Following prebiotic intake, gut microbiome changes were seen, including a notable increase in the relative abundance of fermenters such as Bifidobacterium and Faecalibacterium. Treatment was also associated with changes in bacterial functional pathways associated with either improved energy metabolism (upregulation of tyrosine metabolism) or anti-inflammatory effects (reduced geraniol degradation). There were no differences in stool SCFA levels. Plasma metabolites associated with improved glycemia, such as hippurate, were significantly increased after treatment and there were positive and significant changes in the immune regulatory function of mucosal associated invariant T cells. There was a significant decrease in the area under the curve glucose but not C-peptide, as measured during a mixed meal tolerance testing, following the prebiotic consumption. In summary, the prebiotic HAMS-AB was safe in adolescents with T1D and showed promising effects on the gut microbiome composition, function and immune regulatory function.

## Introduction

Compared to healthy individuals, people with type 1 diabetes (T1D) demonstrate gut dysbiosis [1]. The gut microbiome can be altered using high-amylose maize starch (HAMS), a well-tolerated source of dietary fiber. Following colonic bacterial fermentation, acetylated and butyrylated HAMS (HAMS-AB) releases large amounts of short chain fatty acids (SCFAs) [2], which are anti-inflammatory and immunomodulatory [2, 3].

The main objective of this pilot study was to assess the safety of HAMS-AB and its effect on the gut microbiome in people with recently diagnosed T1D. We used a cross-over design to allow for assessment of prebiotic efficacy through comparison of individuals to themselves as their own controls. Secondary outcomes included HAMS-AB’s effects on glycemia and β-cell function. The full study protocol has been previously published [4]. Briefly, participants were randomized to start with either the prebiotic and a standard ADA recommended diabetes diet at home for 4 weeks or just the diabetes diet for 4 weeks, with a 4-week washout period and then a cross-over to the other arm for 4 weeks (12-week study period).

## Results

Recruitment was from July 2020 to December 2022. Twelve participants were enrolled; 7 finished the study. Three withdrew prior to consumption of HAMS-AB; anxiety around blood draws, family stress and struggling to follow the diabetic diet were the reasons reported. The other two did not tolerate the prebiotic; one developed gagging with attempted consumption, the second developed nausea. Symptoms resolved with prebiotic discontinuation.

Data from the remaining 7 individuals was considered sufficient to proceed to a Phase Ib trial, thus resulting in closure of this Phase Ia trial. Supplemental Table 1 shows baseline characteristics for the 7 participants who completed.

### Safety

Overall, HAMS-AB consumption was safe. Supplemental Table 2 lists the adverse events (AEs) documented. There were no severe adverse events during the trial and all AEs resolved without sequelae. As expected, a few participants developed gastrointestinal symptoms due to increased fiber intake.

### Gut microbiome profile and functional pathway changes

There were no differences seen in alpha diversity measures by treatment period. There were shifts seen in the gut microbiome profile. **Figure 1** shows changes in the relative abundance of several taxa following each treatment period. Notable were an increased relative abundance of Bifidobacterium at the end of prebiotic consumption (p>0.05). Both diet and prebiotic intake decreased the relative abundance of Blautia, which was more prominent at the end of prebiotic consumption (p=0.0057). There was an increase in Faecalibacterium in those who started with a diabetic diet then later changed to prebiotic treatment. The relative abundance of Dialister remained stable after prebiotic consumption compared to a decrease at the end of the diabetic diet. Lastly, Dorea decreased at the end of the each of each period, but more significantly following prebiotic consumption (p=0.007).

**Figure 1:**
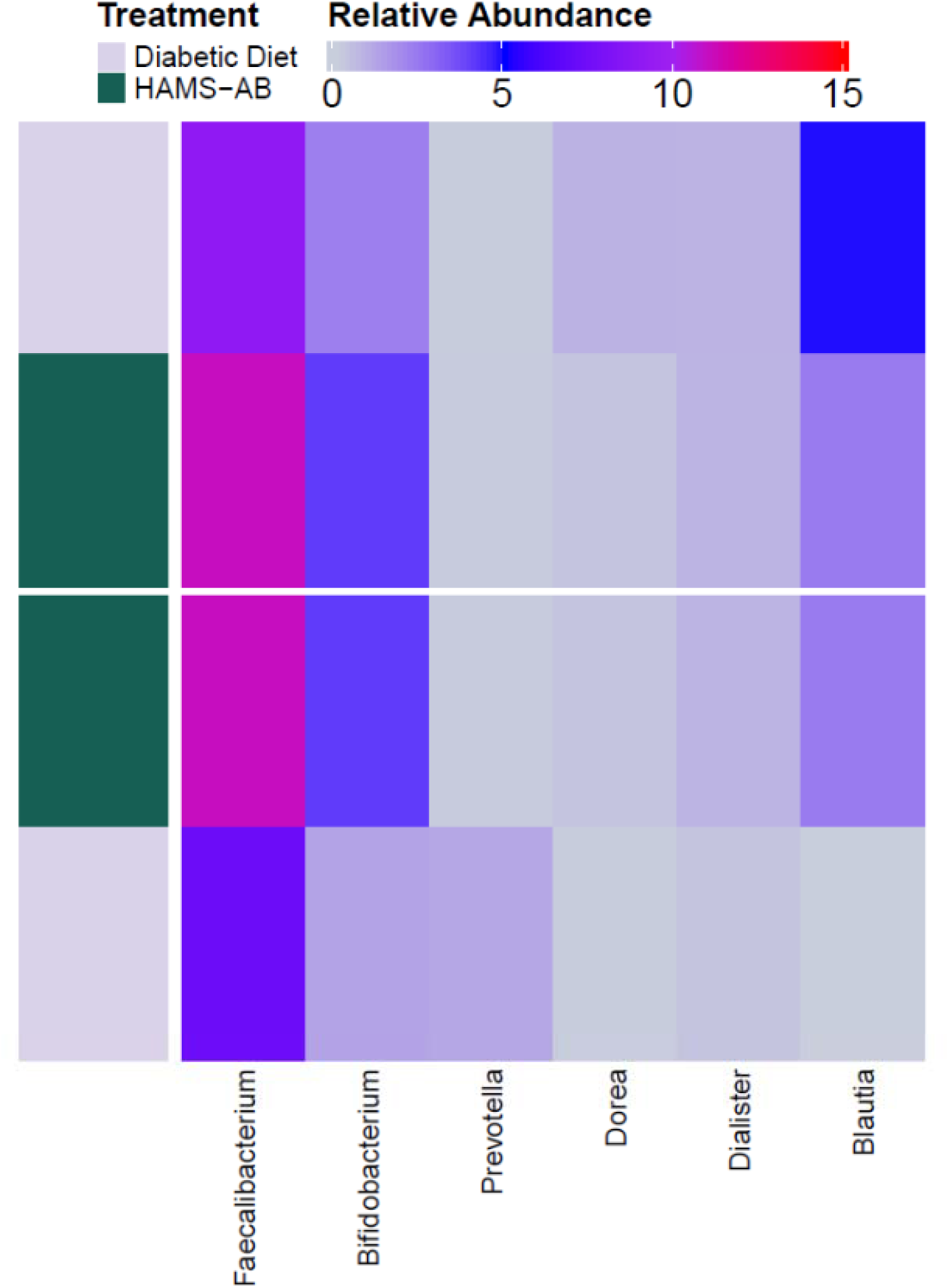
Changes in relative abundance of different species in relation to the intervention type (HAMS-AB vs Diabetic Diet)

Functional pathway analysis showed an upregulation in tyrosine metabolism and a downregulation of Geraniol degradation post-prebiotic consumption.

### Metabolites Analysis

Four weeks post-prebiotic, there was an increase in stool butyrate levels as measured by gas chromatography, from 14.3 ± 6.8 to 23.1 ± 5.6 mmol / kg fecal material (p=0.057). There were smaller increases in acetate or propionate levels (supplemental table 3).

An untargeted plasma metabolomics analysis revealed increases in plasma Hippurate, L-glutamate, tryptophan, and dihydroxyquinoline post-prebiotic consumption (p=0.02 for all after adjusting for multiple comparisons).

### MAIT cells analysis

Mucosal-associated invariant T (MAIT) cells are innate-like T lymphocytes that are activated by bacterial riboflavin. MAIT cells are altered in children at risk for and with T1D [5]. Consumption of HAMS-AB did not change the overall MAIT cell frequency. We saw a reduction in the activation state of MAIT cells post-prebiotic intake, but not in post-diet, marked by reduced CCR6+, CD25+, Granzyme B+ and PD1+ MAIT cells, **Figure 2**.

**Figure 2:**
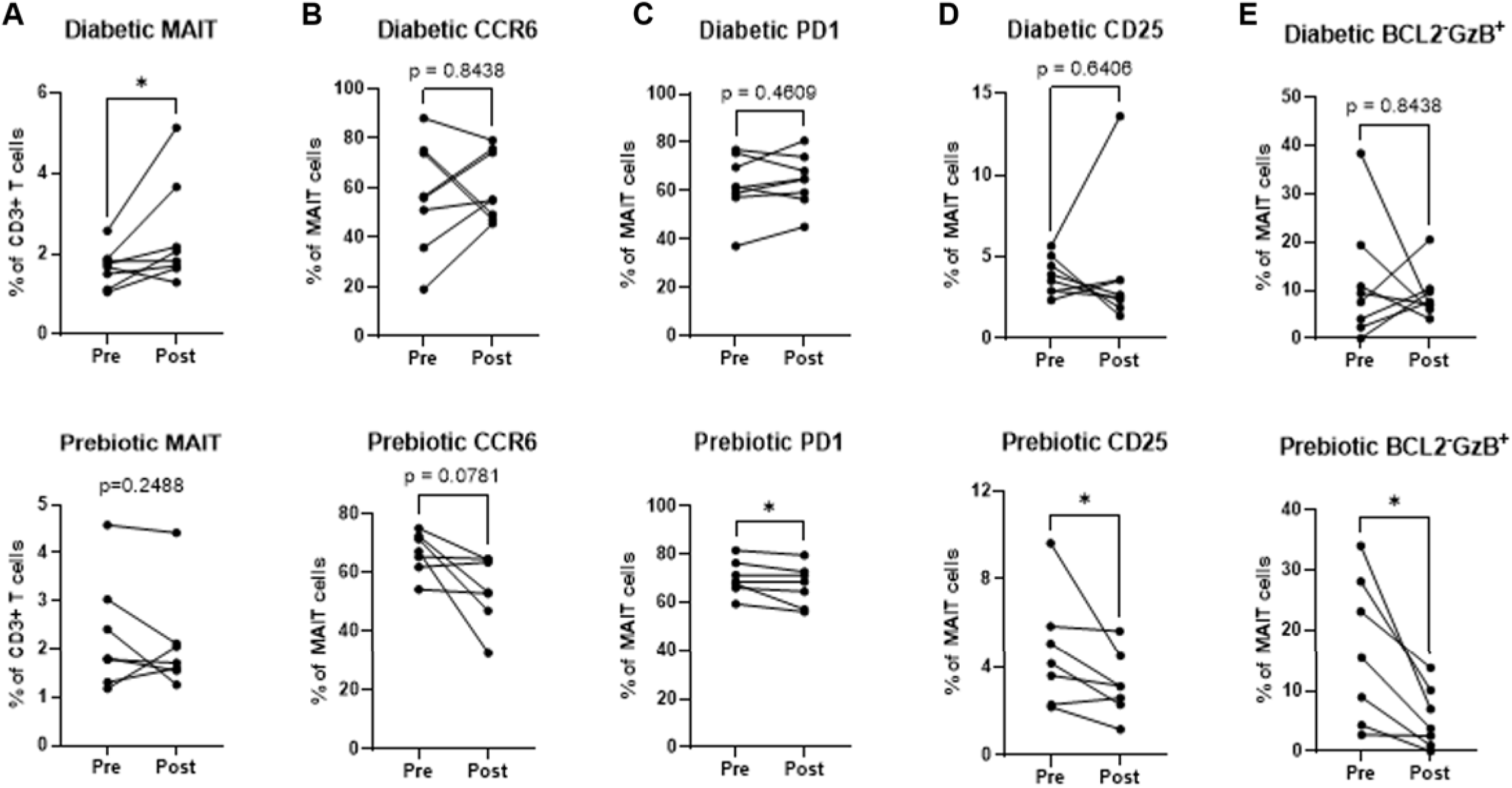
Modulation of the immune response by HAMS-AB. PBMCs were harvested from patients’ blood and activated by MAIT cell antigen 5-OP-RU (A &B) or PMA/Ionomycin (C-E) overnight. MAIT cell phenotypes were analyzed by flow cytometry. Data were analyzed comparing pre- and post- diabetic (upper panels) and prebiotic (lower panels) diets. *, P<0.05, Wilcoxon paired test.

### β cell function

Area under the curve (AUC) measurements from mixed meal tolerance tests (MMTTs) showed that HAMS-AB significantly reduced glucose levels compared to baseline values. An improvement in C-peptide was seen, albeit non-significant (supplemental Table 4).

## Discussion

In this Phase Ia clinical trial, we examined the safety of HAMS-AB consumption in recently diagnosed youths with T1D and its effects on the gut microbiome, metabolites, immune markers and glycemia. We saw an acceptable safety profile of HAMS-AB, with no SAEs. Most AEs were mild/moderate, all resolved before the end of the study period. We saw changes in the gut microbiome composition and metabolite profile associated with prebiotic consumption and changes in immune markers, β-cell function and glycemia. Therefore, our findings underscore the potential for HAMS-AB use in T1D management.

HAMS-AB consumption led to an increased relative abundance of Bifidobacterium and Faecalibacterium. Bifidobacterium is a fermenter and SCFA producer that is typically decreased in both T1D [1]. Faecalibacterium prausnitzii is a butyrate producer and hyperglycemia reduces its abundance [6]. Blautia decreased following prebiotic and diabetes diet alone, although more prominent at the end of the prebiotic consumption period. Blautia abundance is inversely associated with visceral fat adiposity [7]. Lastly, an increase in Dorea and a decrease in Dialister is seen in patients with T1D and their siblings [8], in contrast to what we saw in response to prebiotic.

Overall, SCFAs increased post-prebiotic with a trend towards significance for butyrate. We saw an upregulation in tyrosine metabolism and downregulation of geraniol degradation functional pathways. Tyrosine is an aromatic amino acid that plays an important role in energy metabolism [9], suggesting improved energy metabolism. While geraniol is an acyclic monoterpene alcohol with well-known anti-inflammatory and antimicrobial properties [10]. Therefore, reduced degradation suggests persistence of its anti-inflammatory effects.

Metabolomics analysis revealed an increase in metabolites associated with the gut microbiome, glycemia and energy homeostasis. Hippurate increased post prebiotic and is a microbial metabolite associated with increased gut bacterial diversity and improved glycemia [11]. L-glutamic acid is an important intermediate in metabolism and has been touted with potential for glycemic control [12]. Dihydroxyquinoline has protective and homeostatic effects on the intestinal tract by suppressing inflammation [13]. While tryptophan, partially produced by the gut microbiome, is associated with reduced inflammation and regulating energy homeostasis [14].

MAIT cells are innate-like T cells that are involved in the mucosal immune response [5]. They are thought to play a key role in maintenance of gut integrity, thereby potentially providing a link between the gut microbiome changes and autoimmunity. We saw a reduction in the activation state of MAIT cells marked by reduced CCR6+, CD25+, and PD1+ MAIT cells. This is closely linked to a reduced inflammatory response and promotion of a more regulated immune profile.

This study has strengths. Several measures and metabolic markers were assessed as part of this study. The cross-over design where individuals were their own controls strongly suggests that changes seen were not due to chance alone. Additionally, despite the dropout rate, several measures indicate a positive effect of HAMS-AB. Further, HAMS is a natural supplement that may be favored by many patients. Limitations include the small sample size and short duration of intake. A larger Phase Ib trial to assess these effects is underway (NCT06057454). Further, two individuals did not tolerate the prebiotic, which stresses the individual differences in tolerance to dietary agents and the need for personalized treatment approaches. However, our initial data is encouraging.

## Data Availability

Most data produced in the present work are contained in the manuscript with additional data available upon reasonable requests to the authors.

## Acknowledgements

This study received support from the National Institutes of Health, National Center for Advancing Translational Sciences, Clinical and Translational Sciences Award, Grant Numbers, KL2TR002530 (A Carroll, PI), and UL1TR002529 (A. Shekhar, PI). We also acknowledge support from the Board of Directors of the Indiana University Health Values Fund for Research Award and the Indiana Clinical and Translational Sciences Institute funded, in part by Grant U54TR002529 from the National Institutes of Health, National Center for Advancing Translational Sciences, Clinical and Translational Sciences Award; the Indiana Clinical and Translational Sciences Institute funded, in part by Award Number ULITR002529 from the National Institutes of Health, National Center for Advancing Translational Sciences, Clinical and Translational Sciences Award; the Pilot and Feasibility Grant from the Indiana Center for Diabetes and Metabolic Diseases (P30DK097512); the National Institute Of Diabetes And Digestive And Kidney Diseases of the National Institutes of Health under Award Number K23DK129799, the Doris Duke Charitable Foundation through the COVID□19 Fund to Retain Clinical Scientists Collaborative Grant Program (Grant 2021258) and The John Templeton Foundation (Grant 62288). The content is solely the responsibility of the authors and does not necessarily represent the official views of the National Institutes of Health or other funding agencies.

## Contribution Statement

HMI conceived the study, drafted and edited the manuscript. CEM, LAD, MN and JL contributed to the study design, critically reviewed the manuscript and approved the final version. All have consented to the manuscript publication.

## Data Availability

The data generated from this study will be made available on a case by case basis.

