## Supplementary material for "Safety and effects of acetylated and butyrylated high amylose maize starch in recently diagnosed youths with type 1 diabetes; a Pilot Study": Suppl table and methods

Supplemental Table 1. Demographics for 7 participants who completed the study.

| Mean±SD age (years) | 15.0±1.2 |
| --- | --- |
| Mean±SD diabetes duration (months) | 19.5±6.3 |
| Sex | 5/7 females |
| Race | 4/7 White |
| BMI | <85% |

Supplemental Table 2. Summary of Adverse Events

|  | **Intervention Group** | | **Control Group** | |
| --- | --- | --- | --- | --- |
|  | Affected / at Risk (%) | # Events | Affected / at Risk (%) | # Events |
| Total | 7/9 (77.78%) |  | 7/10 (70%) |  |
| Gastrointestinal disorders | | | | |
| Nausea | 1/9 (11.11%) | 1 | 0/10 (0%) | 0 |
| abdominal pain | 1/9 (11.11%) | 1 | 0/10 (0%) | 0 |
| constipation | 0/9 (0%) | 0 | 2/10 (20%) | 2 |
| emesis | 1/9 (11.11%) | 1 | 0/10 (0%) | 0 |
| flatulence | 1/9 (11.11%) | 1 | 0/10 (0%) | 0 |
| loss of appetite | 0/9 (0%) | 0 | 1/10 (10%) | 1 |
| Infections and infestations | | | | |
| Bilateral halux infection | 0/9 (0%) | 0 | 1/10 (10%) | 1 |
| Cold | 1/9 (11.11%) | 1 | 0/10 (0%) | 0 |
| Yeast infection | 1/9 (11.11%) | 1 | 0/10 (0%) | 0 |
| Investigations | | | | |
| Anxiety | 0/9 (0%) | 0 | 1/10 (10%) | 1 |
| Sweating during blood draw | 0/9 (0%) | 0 | 1/10 (10%) | 3 |
| Metabolism and nutrition disorders | | | | |
| low vitamin D | 0/9 (0%) | 0 | 1/10 (10%) | 1 |
| Musculoskeletal and connective tissue disorders | | | | |
| Dental Pain | 1/9 (11.11%) | 1 | 0/10 (0%) | 0 |
| Nervous system disorders | | | | |
| Headaches | 0/9 (0%) | 0 | 1/10 (10%) | 1 |
| Product Issues | | | | |
| CGM fell off | 0/9 (0%) | 0 | 1/10 (10%) | 1 |
| Respiratory, thoracic and mediastinal disorders | | | | |
| cough | 0/9 (0%) | 0 | 1/10 (10%) | 1 |
| Skin and subcutaneous tissue disorders | | | | |
| rash at sensor insertion site | 0/9 (0%) | 0 | 1/10 (10%) | 1 |
| Vascular disorders | | | | |
| epistaxis | 0/9 (0%) | 0 | 1/10 (10%) | 1 |

**Supplemental Table 3: Changes in SCFAs after 4 weeks of HAMS-AB vs after 4 weeks of Diabetic diet alone.**

|  | Mean (±SD) Butyric Acid | Acetic Acid Mean (±SD) | Propionic Acid Mean (±SD) |
| --- | --- | --- | --- |
| Before Prebiotic | 14.3 (6.8) | 54.4 (25.8) | 14.9 (7.1) |
| After Prebiotic | 23.1 (5.6) | 59.6 (23.1) | 16.0 (8.5) |
| Before Diabetic Diet | 15.0 (12.7) | 58.2 (22.0) | 14.4 (2.9) |
| After Diabetic Diet | 19.0 (5.2) | 55.5 (17.6) | 15.6 (5.1) |

**Supplemental Table 4: Changes in Mixed Meal Tolerance Glucose and C-peptide Values calculated using the Trapezoidal rule. NS indicates non-significant changes.**

|  | | **Baseline** | | | **4-weeks of HAMS-AB (or diet)** | | | |
| --- | --- | --- | --- | --- | --- | --- | --- | --- |
| **Intervention** | **Measure** | **Mean** | **SD** | **N** | **Mean** | **SD** | **N** | **p-value** |
| **HAMS-AB** | **C-peptide (ng/ml)** | **1.96** | **0.31** | **4** | **1.68** | **0.28** | **4** | **NS** |
| **Diet** | **C-peptide (ng/ml)** | **2.71** | **0.55** | **3** | **2.61** | **0.4** | **3** | **NS** |
| **HAMS-AB** | **Glucose (mg/dl)** | **255.93** | **18.93** | **5** | **227.58** | **12.87** | **3** | **0.04** |
| **Diet** | **Glucose (mg/dl)** | **232.16** | **30.56** | **3** | **217.13** | **26.71** | **3** | **NS** |

**Supplemental Methods:**

Additional details regarding the study protocol and methods used for the analysis are outlined in (Ismail, HM et al, 2023).

Metagenomic Sequencing

Metagenomics shotgun sequencing was performed by EzBiome (Gaithersburg, MD, USA). Concentration of genomic DNA was measured using the Qubit Fluorometer dsDNA DNA quantification System (ThermoFisher, USA). 50ng-1ug of genomic DNA was used for library construction using NEBNext® Ultra™ II FS DNA Library Prep Kit for Illumina. Briefly, gDNA was enzymatically sheared, DNA fragment ends were repaired, 3’ adenylated, and ligated to adapters according to the manufacturer’s instructions. The resulting adapter-ligated libraries were PCR-amplified using the following protocol: an initial denaturation step performed at 98°C for 45s followed by 5 cycles of denaturation (98°C, 15 s), annealing (60°C, 30 s) and extension (72°C, 30 sec), and a final elongation of 1 min at 72°C. PCR product was cleaned up from the reaction mix with Mag-Bind RxnPure Plus magnetic beads (Omega Bio-tek, Norcross, GA). The libraries were quantified and qualified using the D1000 ScreenTape on an Agilent 2200 TapeStation instrument. The libraries were normalized and pooled for multiplexed sequencing on an Illumina HiseqX10 sequencer (Illumina, San Diego, CA, USA) using the pair-end 150bp run format.

Metagenomic Taxonomic Profiling

The profiling process started by surveying the potential presence of bacterial and archaeal species for each raw metagenomic sample read by using Kraken2 (Wood, Lu, & Langmead, 2019) and a pre-built core gene database (Chalita et al., 2020) containing k-mers (k=35) of reference genomes obtained from the EzBioCloud database (Yoon et al., 2019). Fungi and Viral full genomes from NCBI’s refseq (https://www.ncbi.nlm.nih.gov/refseq/) were also added to the Kraken2 database. After acquiring a list of candidate species, a custom bowtie2 (Langmead & Salzberg, 2012) database was built utilizing the core genes and genomes from the species found during the first step. The raw sample was then mapped against the bowtie2 database using the --very-sensitive option and a quality threshold of phred33. Samtools (Li et al., 2009) was used to convert and sort the output bam file. Coverage of the mapped reads against the bam file was obtained using Bedtools (Quinlan & Hall, 2010). Then, to avoid false positives, using an in-house script, we quantified all the reads that mapped to a given species only if the total coverage of their core genes (archaea, bacteria) or genome (fungi, virus) was at least 25%. Finally, species abundance was calculated using the total number of reads counted and normalized species abundance was calculated by using the total length of all their references.

Metagenomic Functional Profiling

For each sample, functional annotations were obtained by matching each read, using DIAMOND (Buchfink, Xie, & Huson, 2014), against the KEGG database (Kanehisa, Furumichi, Tanabe, Sato, & Morishima, 2017). DIAMOND was executed using the blastx parameter, which converts each metagenomic read into multiple amino acid sequences by generating all six open reading frame variations, and then matches it against the pre-built KEGG database. If a read had multiple KEGG hits, the top hit was always used. After quantifying all the KEGG orthologs present, minpath (Ye & Doak, 2009) was used to predict the presence of KEGG functional pathways.

Alpha Diversity

All sample read counts were normalized with DESeq2’s median of ratios method (Love et al., 2014. Alpha diversity measures were calculated from normalized taxa counts using the microbiome package in R (Leo Lahti et al, 2012).

Statistical analysis

The Wilcoxon rank sum test between unpaired samples using the ranksums function from the scipy Python package (Virtanen et al. 2020). Linear mixed model analysis using the function lmer from the lme4 R package with the following formula: lmer(var,"~ Treatment + Period + Sequence + (1|ID), data = data) (Bates et al., 2015). Where ‘ID’ is the subject identifier and ‘var’ is the count data of the genus, species, or pathway of interest. Given the cross-over design, a general linear mixed model was used to assess the effect of prebiotics on the gut microbiome. The model formula included the following: Selected Taxa ~ Treatment + Time + Sequence + Period + Treatment*Time (1|ID). Coefficient trends were assessed, and depending on which treatment the participants were first randomized to, period 1 and period 2 were interpreted as follows:

- The Period 1 value represents the relative abundance of the given taxa at the end of the Period for a given treatment sequence.
- Likewise, the Period 2 value represents the relative abundance of the given taxa at the end of Period 2.
- For a given line, the second treatment type at the end of a given treatment sequence (‘Diet’ in the case of Prebiotic -> Diet) is responsible for the trend seen in the slope of the line.

Flow cytometry

PBMCs were harvested from blood draw during each visit. Cells were cryopreserved and stored in liquid nitrogen tank till analyses. Cells were cultured in the presence of either PMA (25 ng/ml) and Ionomycin (1 ug/ml), or 5-OP-RU (MeG 25 uM, 5-A-RU 1 uM) overnight. Brefeldin A was added 3 hours before harvest. All cells were firstly surface-stained using antibodies specific for the following markers:CD3, CD4, CD8, TCR Va7.2, CD161, MR1 tetramer, CD25, CD56, CD69, PD-1 and CCR6. These cells were fixed, permeabilized using and stained for intracellular and nuclear markers: IFNg, TNFa, IL-17A, BCL-2, Granzyme B. Flow data were acquired using Cytek spectral flow cytometer and analyzed using FlowJo.

| **Antibody name** | **Company** | **Catalog #** |
| --- | --- | --- |
| CD69-PE/Dzaale594 | Biolegend | 310941 |
| CD56-BV510 | Biolegend | 318339 |
| CCR6-APC/Cy7 | Biolegend | 353431 |
| BCL-2-BV421 | Biolegend | 658709 |
| IFNg-BV605 | Biolegend | 506541 |
| TNFa-BV785 | Biolegend | 502947 |
| GzB-PE/Dazzle | Biolegend | 372215 |
| IL-17A-BV570 | Biolegend | 512329 |
| IL-4-APC/Cy7 | Biolegend | 500833 |
| CD161-AF488 | Biolegend | 339924 |
| TCR Va7.2 PE | Biolegend | 351706 |
| MR1 tetramer-APC | NIH Tetramer Core |  |
| CD3-AF700 | BD Biosciences | 557943 |
| CD25-PerCP-Cy5.5 | BD Biosciences | 560503 |
| CD4-Pacific Blue | BD Biosciences | 558116 |
| CD8-PE/Cy7 | BD Biosciences | 566859 |
